# Food insecurity and other barriers to adherence to a gluten-free diet in individuals with celiac disease and non-celiac gluten sensitivity in the Netherlands: a mixed-methods study

**DOI:** 10.1101/2024.04.28.24306418

**Authors:** Sharine M. Smeets, Jessica C. Kiefte-de Jong, Laura A. van der Velde

## Abstract

**Objectives:** To determine the prevalence of food insecurity among individuals with celiac disease (CeD) and non-celiac gluten sensitivity (NCGS) in the Netherlands and explore its association with diet quality and other barriers to adherence to a gluten-free diet.

**Design:** Mixed-method design comprising a survey and semi-structured interviews.

**Setting:** An online survey was distributed through social media accounts and the newsletter of the Dutch Association for Celiac Disease. Community-dwelling patients were surveyed and interviewed between June and November 2023.

**Participants and outcome measures:** In total 548 adults with CeD and NCGS in the Netherlands who adhered to a gluten-free diet completed the survey including questions related to demographics, household food security, financial stress and dietary quality. Regression analyses were conducted to assess associations between food insecurity and diet quality, and between food insecurity and perceived difficulty of gluten-free eating and cooking. Additionally, semi-structured interviews with 8 food insecure adults with CeD were conducted.

**Results:** The prevalence of food insecurity was 23.2%, with 10.4% reporting very low food security. Very low insecurity was associated with poorer diet quality (β=-5.5; 95%CI=-9.2,-1.9; p=0.003). Food insecurity was associated with heightened perceived barriers across multiple themes, including skills, social circumstances, resources and gluten-free products, with odds ratios ranging between 1.9-4.7 for crude models (very low food security vs food security). The qualitative analysis provided a deeper understanding of these challenges, including employed strategies to manage costs and insights into the mental burden associated with adhering to a gluten-free diet.

**Conclusion:** These findings indicate that food insecurity is prevalent among Dutch people with CeD and NCGS, with potential impact on dietary quality and adherence to a gluten-free diet. It further provided insight into perceived barriers to adhering to a gluten-free diet among this target population. These challenges should be taken into account by clinicians and policy makers.

**STRENGTHS AND LIMITATIONS:**

- The study provides previously unexplored insights in food insecurity among people with CeD and NCGS in the Netherlands.
- By employing a mixed-methods study design, the quantitative findings gained added depth as they were enriched by the personal experiences elucidated in the qualitative analysis. These provided a richer understanding of the challenges individuals face, including employed strategies to manage costs and insights into the mental burden associated with adhering to a GF diet.
- An inherent limitation of the study is its cross-sectional design, which prevents drawing causal conclusions regarding factors associated with food insecurity.
- The study relied on self-reported data through anonymous surveys, which introduces the possibility of recall bias and social desirability bias
- In the context of this study, food insecurity might be limited to ‘gluten-free food insecurity’ in otherwise food secure households.

## INTRODUCTION

Food insecurity remains an important global issue, both in developing nations and in high-income countries. The prevalence of food insecurity in Northern America and Europe has increased to 8.0% in 2022. Food security can be defined as having “physical and economic access to sufficient, safe and nutritious food to meet dietary needs and food preferences for an active and healthy life.”(1) Whereas in the past research on food security was mostly focused on developing countries, there has been an increase of food security research in high-income countries in recent years(2). This was intensified amidst the COVID-19 pandemic, with worldwide food shortages, increased food prices and loss of income making food insecurity more visible(3). Part of the research on food insecurity has focused on target populations that appear to be at risk for food insecurity, such as low-income households, ethnic minorities and senior citizens(4–9). In the Netherlands, limited food security research has been done, but it has identified a 73% prevalence of food insecurity among food bank recipients and a 26% prevalence in disadvantaged neighborhoods(10, 11).

Individuals with medical dietary restrictions, such as food allergies or hypersensitivities, may be another group at risk for food insecurity, as they face diets that are often expensive and difficult to access(12). Few studies have been conducted on food insecurity among people with food hypersensitivity and specifically celiac disease (CeD). CeD is an auto-immune disorder which primarily affects the small intestine and is triggered by the ingestion of gluten. While there is active research into pharmacological therapy, current treatment consists of adhering to a gluten-free diet(13). Along with people with CeD, people with non-celiac gluten sensitivity (NCGS) also benefit from following a gluten-free diet. These are people without CeD or wheat hypersensitivity who suffer from reproducible symptomatic responses to gluten-containing foods(14).

However, adhering to a gluten-free diet can be a challenge. Compared to their gluten-containing counterparts, gluten-free products can be up to four times more expensive(15, 16). Therefore, adhering to a gluten-free diet is associated with an increased economic burden for people with CeD or NCGS(15). This economic burden is expected to be even higher at present, as food prices in the European Union have risen dramatically since the beginning of 2022(17). In addition to costs, the availability of gluten-free products and inadequate food labeling have been identified previously as major barriers to following a gluten-free diet(18). Furthermore, eating out at restaurants or attending social events may also pose a challenge, due to the limited availability of gluten-free food options. Aside from these obstacles, factors influencing the belief in one’s ability to adhere to a gluten-free diet, such as concerns about availability of gluten-free foods, may also play a role in gluten-free diet adherence. These fall within the realm of ‘perceived behavioral control’ within the theory of planned behavior (TPB)(19). Prior TPB research among people with CeD has shown that perceived behavioral control, as well as one’s ‘attitude’ toward the diet, significantly influences overall adherence to a gluten-free diet(20, 21). This could imply that individuals experiencing food insecurity might encounter increased challenges in adhering to a gluten-free diet, given that their financial situation could negatively impact their physical and economic access to gluten-free products as well as their beliefs in being able to adhere to the diet, which may also result in a less positive attitude towards the diet.

Prior research on food security among people with CeD has shown an increase of food insecurity among CeD households in the United States during the COVID-19 pandemic(22, 23). A recent review of national survey data collected in the United States between 2008 and 2014, revealed the negative effects of food insecurity in individuals with CeD. Food insecurity was found to be significantly associated with decreased adherence to a gluten-free diet, as well as a reduced intake of essential nutrients, such as protein, various vitamins and several minerals(24). Notably, individuals with CeD already are at higher risk of certain vitamin and mineral deficiencies due to malabsorption, making them particularly vulnerable to the adverse effects of food insecurity. This raises concerns about potential health issues, such as anemia and low bone density(25). Building upon research conducted in the United States on food insecurity among people with CeD, this study aims to provide an insight into the situation in the Netherlands. The objectives of this study are to determine the prevalence of food insecurity among Dutch individuals with CeD and NCGS and to identify potential associations between food insecurity, dietary intake and perceived barriers to adhering to a gluten-free diet.

## METHODS

### Study design and data collection

This study employed a cross-sectional design to survey adults with CeD or NCGS in the Netherlands. First, an anonymous electronic survey was distributed between June and September 2023 through social media accounts (Facebook and Instagram) and the newsletter of the Dutch Association for Celiac Disease. The questionnaire contained several sub-sections, each addressing different aspects: sociodemographic and lifestyle factors, participant CeD or NCGS status, food insecurity and experienced financial scarcity, dietary intake, experiences with adhering to gluten-free diet (including open questions) and self-reported mental wellbeing. The full questionnaire is available in Additional Document 1. The questionnaire was available to participants aged ≥18 years and in Dutch only. There were a total of 987 survey responses. Incomplete survey responses, responses from participants not dealing with CeD or NCGS and responses from participants not following a gluten-free diet were excluded from the analysis. This resulted in 548 completed questionnaires (Figure 1).

**Figure 1.**
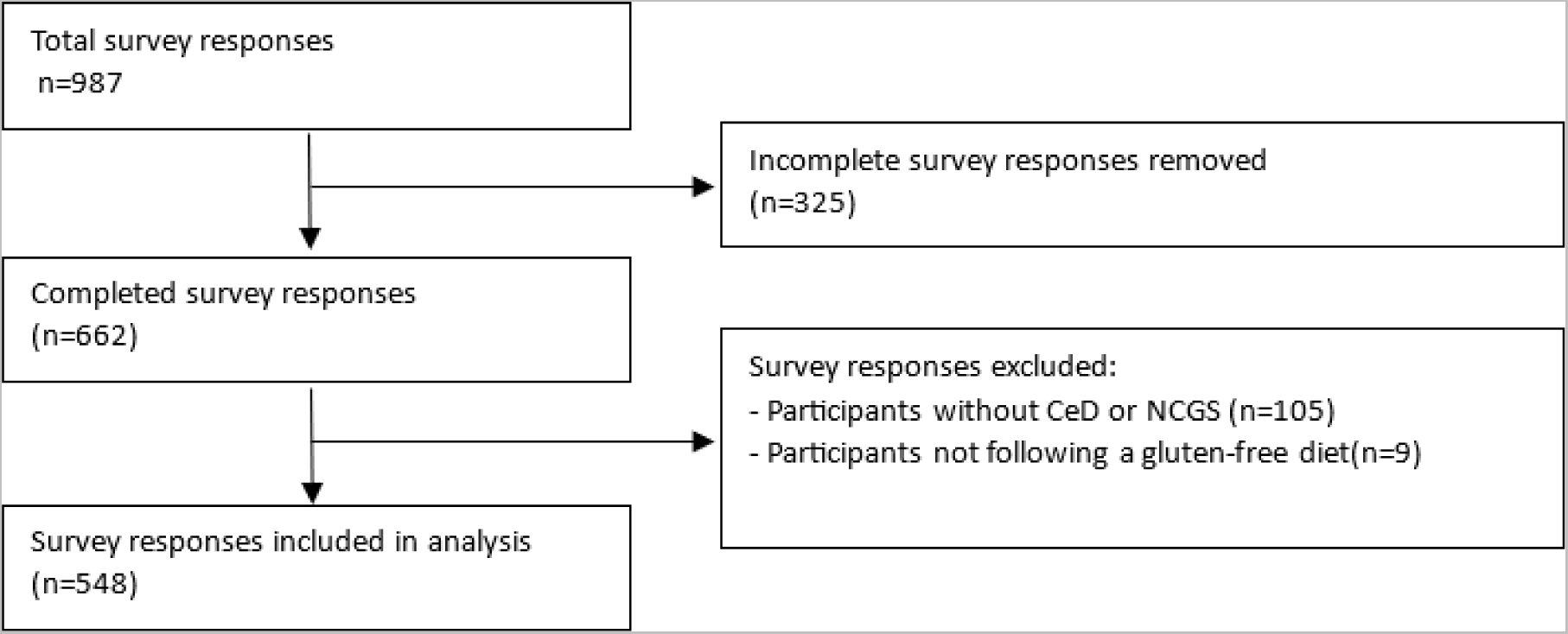
Study participants flow diagram.

Secondly, we conducted a qualitative exploration through interviews of persons with CeD or NCGS in November 2023, employing an inductive analysis approach. For this, new participants were recruited through social media accounts of the Dutch Association for Celiac Disease. A convenience sample of eleven participants experiencing food insecurity, taking into account the diversity of the sample, were approached. Three participants were unable reach after initial information was provided. In total, eight participants were interviewed. Informed consent using a digital consent form was obtained from all participants. None of the participants who agreed to participate dropped out of the study.

The study was reviewed by the Medical Ethics Committee of Leiden University Medical Centre (LUMC) and confirmed not to be subject to the Medical Research Involving Human Subjects Act (WMO) (P17.164).

### Survey design and measures

Self-reported sociodemographic information and lifestyle factors were collected in the survey. Sociodemographic data included age, sex, household size and composition, presence of other people with CeD or NCGS in the household, current employment, net monthly income, educational level and ethnicity. Personal net monthly income was presented in three categories: less than €1500, between €1500-€2500 and more than €2500. The educational level was classified based on the International Standard Classification of Education(26) into a lower educational level (ISCED ≤3, up to and including upper secondary education) and higher educational level (ISCED ≥4, from post-secondary non-tertiary education). Ethnicity was determined by asking “Which ethnicity do you feel most connected to?” and was categorized as either having a Dutch or non-Dutch ethnicity. Lifestyle factors included height, weight and smoking status (yes/no/in the past). Body Mass Index (BMI, kg/m2) was calculated using the self-reported weight and height. Subsequently, the BMI values were categorized as follows: underweight (BMI < 18.5 kg/m2), normal weight (BMI 18.5–25 kg/m2), overweight (BMI 25–30 kg/m2), and obese (BMI ≥ 30 kg/m2), based on the WHO cut-off points(27).

The celiac disease status of participants was determined with the question: “Do you, or someone in your household or someone other close to you have celiac disease? If so, who?” Participants were allowed to provide multiple answers. If at least “Yes, I have celiac disease myself” was answered affirmatively, the participant was considered to have celiac disease. For NCGS participant status, the identical question was presented, with ‘celiac disease’ being substituted with ‘gluten sensitivity’. If a participant answered both the CeD question and the NCGS question affirmatively, they were categorized as a CeD participant.

Food security status was assessed using the USDA 6-item Short Form of the Food Security Survey Module(28), translated to Dutch. ‘(Gluten-free)’ was added in each prompt before ‘food’ or ‘meals’. Food security scores theoretically ranged between 0 and 6 and were presented as two categories: food secure (total score 0-1) and food insecure (total score 2-6). For a subset analysis, they were divided into three categories: high food security (total score 0-1), low food security (total score 2-4) and very low food security (total score 5-6), according to USDA standards(28).

Experienced financial scarcity was assessed using 5 selected statements of the 12-item Psychological Inventory of Financial Scarcity (PIFS), translated to Dutch(29). The following statements were included: ‘I often don’t have enough money’, ‘ I am constantly wondering whether I have enough money’, ‘I often worry about money’, ‘I am only focusing on what I have to pay at this moment rather than my future expenses’ and ‘ I experience little control over my financial situation’. For each statement, the score ranged between a minimum of 1 and maximum score of 5, resulting in a combined score with a theoretical minimum of 5 and maximum of 25, with higher scores indicating higher experienced financial scarcity.

Self-reported mental well-being was assessed using the WHO well-being index (WHO-5)(30), translated to Dutch. The raw score ranged from 0 to 25. A percentage score ranging from 0 to 100 was presented by multiplying the raw score by 4, with 0 representing the worst possible mental well-being and 100 representing the best mental well-being, according to standards(30).

Diet quality was assessed using a short food frequency questionnaire (FFQ), based on current Dutch national dietary intake recommendations from the Dutch Health Council (DHC) and the Netherlands Nutrition Center (NNC) (31, 32). The FFQ consisted of questions about 13 food group components, including vegetables, fruit, legumes, unsalted nuts, fish, (gluten free) grain products, dairy, tea, coffee, oils and fats, sugar-containing beverages, savory and sweet snacks. For each component, a minimum score of 0 and a maximum score of 10 could be obtained, resulting in a total diet score ranging from a theoretical minimum of 0 to a theoretical maximum of 130, with higher scores indicating better adherence to the dietary guidelines. If data were missing for a dietary component, the least favorable outcome was assumed. This occurred in 102 cases (19%) for one dietary component, 17 cases (3%) for two dietary components and in 2 cases for three and four dietary components (0,8%). Detailed information of the dietary intake assessment and diet quality score calculation is provided in Additional Table 1.

**Table 1.** Total food secure and food insecure participants and food security status in three categories.

| Food security status | n (%) |
| --- | --- |
| Food secure (total) | 417 (76,8%) |
| Food insecure (total) | 127 (23,2%) |
| Low food security | 70 (12,8%) |
| Very low food security | 57 (10,4%) |

Perceived barriers to adhering to a gluten-free diet were analyzed using nineteen 5-point Likert scale statements on the perceived difficulty of gluten-free eating and cooking and seven 5-point Likert scale statements on attitude towards gluten-free products, based on the Theory of Planned Behavior(19). The statements included topics such as price, availability, quality, variety, nutrition content and eating out and can be found in the questionnaire provided in Additional Document 1.

### Interview design

Food insecurity status of participants was determined prior to recruitment using the USDA 6-item Short Form of the Food Security Survey Module(28), translated to Dutch. Only participants that where classified as being food insecure were invited to participate in the interview. Open interviews were conducted by telephone and video calls, following a guided topic list developed based on issues raised in the initial questionnaire. The list included various discussion topics and open-ended example questions for each topic to guide the interviewer. Prior to the interviews, these topics and questions were reviewed and discussed by the research team. Interviews started with general inquiries about the participants’ backgrounds and living situations to establish rapport and put the participants at ease. Subsequently, the questions delved into their experiences with adhering to a gluten-free diet, covering aspects such as costs, availability and social implications. The interviewees were encouraged to introduce any additional topics of interest to them during the conversation.

After each interview the topic list was reassessed, and adjustments or new topics were incorporated as necessary, reflecting emerging themes from the discussions. The full interview guide is available in Additional Document 2. During the interviews, only the participant and interviewer were present.

All interviews were audio-recorded using a digital voice recorder or online recording software and transcribed verbatim. Interviews were scheduled at times convenient for the participants. Written informed consent was obtained from all participants. Participants received a €20 gift card for their participation.

### Survey statistical analysis

Statistical analyses were performed using IBM SPSS Statistics v29. Subject and household characteristics were described as mean (SD) for continuous variables and frequencies and percentages for categorical variables. Differences between the food secure and food insecure group were assessed with Mann–Whitney U testing for continuous variables and chi-squared testing for categorical variables. In 4 cases (0.8%), implausible answers were observed. In two cases, these were answers to the question on household size and were corrected based on further information that was provided in the questionnaire. In the two other cases, these were answers to the question on body weight and data was imputed based on the median body weight of other respondents with identical body heights.

An explanatory factor analysis was run on the nineteen 5-point Likert statements about the perceived difficulty of gluten-free eating and cooking. Statements were combined into one component based on the pattern matrix using Direct Oblimin factor rotation with 0.4 as minimal loading. The component scores were calculated by combining the scores of the separate statements and dividing them by the number of statements, resulting in a theoretical minimum score of 1 and maximum score of 5 for each component. For each component, scores were then dichotomized into disagreeing (score 1-3) or agreeing (score 4-5) with the statements. The seven 5-point Likert scale statements on attitude towards gluten-free products were combined into one by adding the scores of the separate statements together and dividing them by the number of statements, resulting in a theoretical minimum score of 1 and maximum score of 5, with higher scores indicating a more positive attitude towards gluten-free products.

Multiple linear regression analysis was conducted to investigate the association between food insecurity and diet quality. The analysis was performed using the continuous total diet quality score as the dependent variable. Two models were presented. For the first (crude) model, food security status was analyzed as an independent variable in three categories: food secure, low food secure and very low food secure. The second model was adjusted for age, educational level and net monthly income.

Logistic regression analyses were conducted to study the association between food insecurity and the perceived difficulty of gluten-free eating and cooking. The analyses were performed with the components resulting from the factor analysis as dependent variables. Two models were presented for each component. For the first (crude) model, food security status was analyzed as an independent variable in three categories: food secure, low food secure and very low food secure.

The second model was adjusted for age, educational level and net monthly income. A two-sided p-value of 0.05 was considered statistically significant for all analyses.

### Qualitative analysis

A general inductive approach was used to analyze the qualitative data. Parts of the interview transcripts were coded using open coding. Some text segments were assigned more than one code category and text segments that were not relevant for the study objectives were not included in any category. Throughout this process, some codes with similar meanings were merged. Codes and code assignment was discussed within the research team. Codes were grouped into themes. The software Atlas.ti v23.2.3 was used to assist with the coding process and extraction of quotes and themes.

### Patient and public involvement

The director and information manager of the Dutch Association for Celiac Disease were involved in participant recruitment and advise regarding the questionnaire and interview content, to ensure that the questions were relevant and understandable.

## RESULTS

### Questionnaire

#### Participant characteristics

In total, 548 participants were included in the analysis. The overall prevalence of food insecurity was 23.2%; 12.8% of participants experienced low food security and 10.4% experienced very low food security (Table 1). Table 2 summarizes the characteristics of the survey respondents for the total population and by food security status. Most participants were female (90.3%) and had celiac disease (86.7%). The mean age of the total study sample was 44.7± 14.6 years. The majority of the participants were of Dutch ethnicity. Compared to food secure (FS) participants, food insecure (FI) participants were often younger, had a lower income and a lower educational level. FI participants had a higher mean financial scarcity score (i.e., had a stronger perceived financial scarcity) and a lower WHO-5 score (i.e., had a poorer perceived mental wellbeing) compared to FS participants.

**Table 2.** Sociodemographic characteristics of participants (total n=548)

| Variable | Total | Food secure | Food insecure | P-value* |
| --- | --- | --- | --- | --- |
| n (%) | 548 (100%) | 421 (76,8%) | 127 (23,2%) |  |
| Age (y) [mean (SD)] | 44,7 (14,6) | 46,1 (14,5) | 40,0 (14,3) | <0.001 |
| Female [n (%)] | 495 (90,3%) | 375 (89,1%) | 120 (94,5%) | 0.183 |
| Person with CeD/NCGS [n (%)] |  |  |  | 0.455 |
| Person with celiac disease | 475 (86,7%) | 363 (86,2%) | 112 (88,2%) |  |
| Person with NCGS | 73 (13,3%) | 58 (13,8%) | 15 (11,8%) |  |
| Household size [mean (SD)] | 2,8 (1,5) | 2,8 (1,5) | 2,7 (1,5) | 0.111 |
| Household composition [n (%)] |  |  |  | 0.608 |
| Single parent | 13 (2,4%) | 9 (2,1%) | 4 (3,1%) |  |
| Multiple household with children | 191 (34,9%) | 151 (35,9%) | 40 (31,5%) |  |
| No children household | 344 (62,8%) | 261 (62,0%) | 83 (65,4%) |  |
| Other household members with CeD/NCGS [Yes, n (%)] | 128 (23,4%) | 97 (23,0%) | 31 (24,4%) | 0.749 |
| Current paid job [Yes, n (%)] | 409 (74,6%) | 321 (76,2%) | 88 (69,3%) | 0.103 |
| Personal net monthly income (€) [n (%)] |  |  |  | <0.001 |
| <1500 | 128 (23,4%) | 76 (18,1%) | 52 (40,9%) |  |
| 1500-2500 | 207 (37,8%) | 156 (37,1%) | 51 (40,2%) |  |
| >2500 | 213 (38,9%) | 189 (44,9%) | 24 (18,9%) |  |
| Educational level [n (%)] |  |  |  | 0.006 |
| Lower | 207 (37,8%) | 146 (34,7%) | 61 (48,0%) |  |
| Higher | 341 (62,2%) | 275 (65,3%) | 66 (52,0%) |  |
| Financial scarcity score [mean (SD)] | 11,6 (4,9) | 10,0 (4,1) | 16,7 (3,6) | <0.001 |
| Ethnicity [n (%)] |  |  |  | 0.957 |
| Dutch | 526 (96,0%) | 404 (96,0%) | 122 (96,1%) |  |
| Non-Dutch | 22 (4,0%) | 17 (4,0%) | 5 (3,9%) |  |
| Weight status |  |  |  | 0.414 |
| Underweight (BMI <18,5) | 20 (3,6%) | 17 (4,0%) | 3 (2,4%) |  |
| Normal weight (BMI 18,5-25) | 304 (55,5%) | 232 (55,1%) | 72 (56,7%) |  |
| Overweight (BMI 25-30) | 143 (26,1) | 114 (27,1%) | 29 (22,8%) |  |
| Obese (BMI >30) | 81 (14,8%) | 58 (13,8%) | 23 (18,1%) |  |
| Smoking status [n (%)] |  |  |  | 0.307 |
| Yes | 15 (2,7%) | 10 (2,4%) | 5 (3,9%) |  |
| No | 470 (85,8%) | 366 (86,9%) | 104 (81,9%) |  |
| In the past | 63 (11,5%) | 45 (10,7%) | 18 (14,2%) |  |
| WHO-5 score [mean (SD)] | 57,7 (19,5) | 59,5 (18,7) | 51,8 (21,2) | <0.001 |

\*P-value of food secure versus food insecure participants

#### Food insecurity and diet quality

The mean diet quality score out of a maximum score of 130 was 74.9±13.0 for FS participants, 72.1±12.6 for low food secure participants and 69.4±15.0 for very low food secure participants (data not shown). Food insecurity was associated with a significantly lower diet quality score compared to FS participants in very low food secure participants (β=-5.5; 95%CI=-9.2, -1.9; p=0.003), but not low food secure participants (β=-2.8; 95%CI= -6.1, 0.6; p=0.105). Controlling for age, income and education, the association remained statistically significant for very low food secure participants (Table 3).

**Table 3.** Associations between food security status and diet quality.

|  | Low food security |  | Very low food security |  |
| --- | --- | --- | --- | --- |
| | $\beta^*$ (95%CI) | P-value | $\beta^*$ (95%CI) | P-value |
| **Crude model** | -2.76 (-6.11, +0.58) | 0.105 | -5.52 (-9.17, -1.86) | <u>0.003</u> |
| **Adjusted for age, income and education** | -0.92 (-4.22, +2.38) | 0.584 | -3.76 (-7.32, -0.20) | <u>0.039</u> |

\* $\beta$ represents the difference in diet quality score compared to food secure participants.

#### Food insecurity and perceived barriers to adherence to a gluten-free diet

##### Perceived difficulty of gluten-free eating and cooking

The factor analysis of statements on the perceived difficulty of gluten-free eating and cooking resulted in 4 components with total initial eigenvalues greater than 1, explaining 67.2% of total variance. Component 1 comprised six statements regarding skills in gluten-free cooking (e.g. healthy, tasteful), which explained 46.1% of the variance with factor loadings from 0.7 to 0.8. Component 2 comprised five statements regarding social circumstances (e.g. with friends and in restaurants), which explained 7.8% of the variance with factor loadings from 0.5 to 0.9. Component 3 comprised six statements regarding resources (e.g. money and time), which explained 6.9% of the variance with factor loadings from 0.6 to 0.8. Component 4 comprised two statements regarding naturally gluten-free products, which explained 6.3% of the variance with factor loadings from 0.9 to 0.9. A comprehensive breakdown of the statements that make up each component can be found in Additional Document 3.

FI participants were significantly more likely to experience difficulty with gluten-free eating and cooking across all four components compared with FS participants (Table 4). In the age, income and education adjusted model, low food secure participants were more likely to experience difficulty regarding skills (OR=2.5; 95%CI=1.5, 4.3; p=<0.001), social circumstances (OR=2.6; 95% CI=1.1, 6.4; p=0.038), resources (OR=2.5; 95% CI=1.5, 4.4; p=0.001) and naturally gluten-free products (OR=1.8; 95%CI=1.0, 3.1; p=0.045) in gluten-free eating and cooking compared to FS participants. In the adjusted model, participants with very low food security were more likely to experience difficulty regarding skills (OR=4.4; 95%CI=2.4, 8.1; p=<0.001) and resources (OR=4.2; 95%CI=2.3, 7.8; p<0.001) in gluten-free eating and cooking as well. Compared to the crude model, the odds ratios were slightly lower in the adjusted model regarding social circumstances and naturally gluten-free products and not statistically significant in very low food secure participants (Table 4).

**Table 4.**
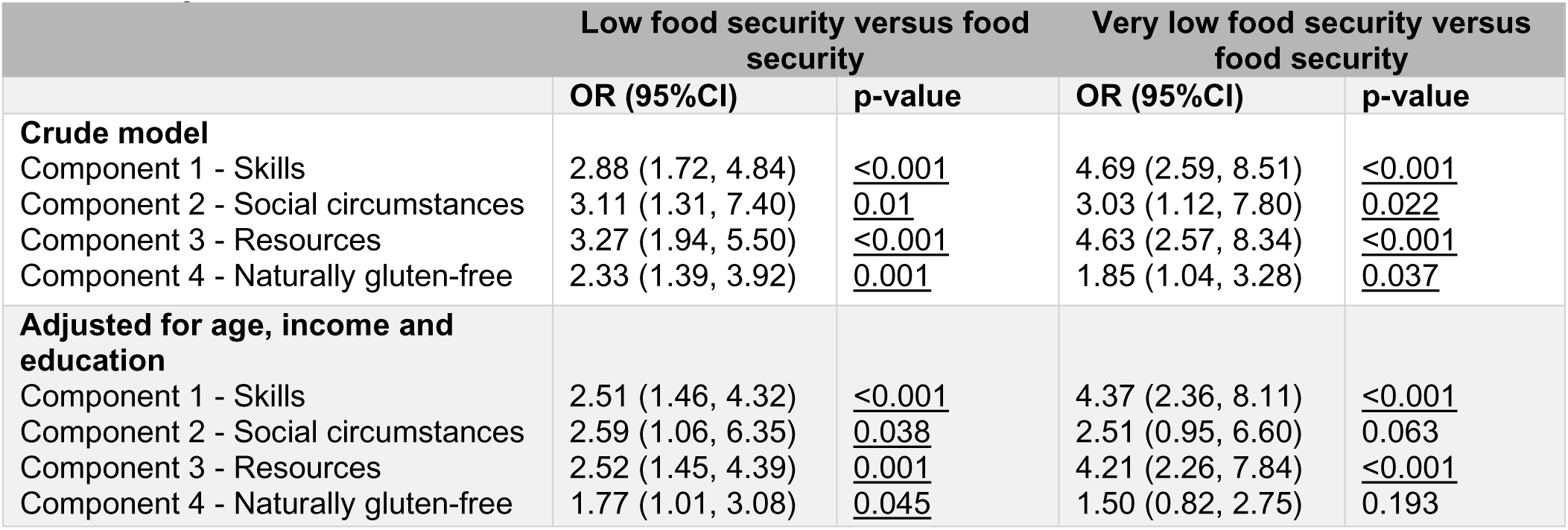
Associations between food security status and perceived difficulty of gluten-free eating and cooking.

|  | Low food security versus food security |  | Very low food security versus food security |  |
| --- | --- | --- | --- | --- |
|  | OR (95%CI) | p-value | OR (95%CI) | p-value |
| **Crude model** |  |  |  |  |
| Component 1 - Skills | 2.88 (1.72, 4.84) | <u><0.001</u> | 4.69 (2.59, 8.51) | <u><0.001</u> |
| Component 2 - Social circumstances | 3.11 (1.31, 7.40) | <u>0.01</u> | 3.03 (1.12, 7.80) | <u>0.022</u> |
| Component 3 - Resources | 3.27 (1.94, 5.50) | <u><0.001</u> | 4.63 (2.57, 8.34) | <u><0.001</u> |
| Component 4 - Naturally gluten-free | 2.33 (1.39, 3.92) | <u>0.001</u> | 1.85 (1.04, 3.28) | <u>0.037</u> |
| **Adjusted for age, income and education** |  |  |  |  |
| Component 1 - Skills | 2.51 (1.46, 4.32) | <u><0.001</u> | 4.37 (2.36, 8.11) | <u><0.001</u> |
| Component 2 - Social circumstances | 2.59 (1.06, 6.35) | <u>0.038</u> | 2.51 (0.95, 6.60) | 0.063 |
| Component 3 - Resources | 2.52 (1.45, 4.39) | <u>0.001</u> | 4.21 (2.26, 7.84) | <u><0.001</u> |
| Component 4 - Naturally gluten-free | 1.77 (1.01, 3.08) | <u>0.045</u> | 1.50 (0.82, 2.75) | 0.193 |

##### Attitude towards gluten-free products

The mean attitude towards gluten-free products, with scores ranging from 1 (most negative attitude) to 5 (most positive attitude), was 2.4±0.6 for FS participants, 2.1±0.5 for low food secure participants and 2.06±0.65 for very low food secure participants (data not shown). Table 5 shows two models regarding the associations between food insecurity and attitude towards gluten-free products. Food insecurity was significantly associated with having a lower attitude score (i.e., a more negative attitude) towards gluten-free products in both models, with similar results in low and very low food secure participants.

**Table 5.** Associations between food security status and attitude towards gluten-free products.

|  | Low food security |  | Very low food security |  |
| --- | --- | --- | --- | --- |
| | $\beta^*$ (95%CI) | p-value | $\beta^*$ (95%CI) | p-value |
| **Crude model** | -0.36 (-0.52, -0.20) | <u><0.001</u> | -0.36 (-0.53, -0.19) | <u><0.001</u> |
| **Adjusted for age, income and education** | -0.32 (-0.48, -0.15) | <u><0.001</u> | -0.34 (-0.51, -0.17) | <u><0.001</u> |

\* $\beta$ represents the difference in attitude score compared to food secure participants.

### Interviews

In total, 8 interviews were conducted, with durations ranging from 40 to 57 minutes, averaging 47 minutes.

#### Participant characteristics

One male and seven females were interviewed, aged between 20 and 57 years. All participants were food insecure, two of whom experienced very low food security. The number of years on the gluten-free diet varied between 1 and 23 years (Table 6).

**Table 6.** Sociodemographic characteristics of participants (n=8)

| Participant number | Age range | Sex | Food security status | Civil status | Years on the gluten-free diet |
| --- | --- | --- | --- | --- | --- |
| 1 | 56-60 | Female | Low food security | Single | 16 |
| 2 | 21-25 | Female | Very low food security | Single | 23 |
| 3 | 21-25 | Female | Low food security | Single | 11 |
| 4 | 16-20 | Male | Low food security | Single | 1 |
| 5 | 26-30 | Female | Low food security | Married / cohabitating | 5 |
| 6 | 56-60 | Female | Low food security | Married / cohabitating | 14 |
| 7 | 36-40 | Female | Very low food security | Single | 15 |
| 8 | 46-50 | Female | Low food security | Married / cohabitating | 8 |

#### Key themes related to perceived barriers to adhering to a gluten-free diet

The participants shared various perceived barriers to adhering to a gluten-free diet, as well as some facilitators, within four overarching emerging themes. The four main themes related to perceived barriers to adhering to a gluten-free diet, along with their corresponding subthemes identified in the analysis, are described below and visually represented in Table 7. Further clarification of these themes is offered through excerpts from the interview transcripts.

**Table 7.** Themes and subthemes associated with perceived barriers to adhering to a GF diet in food insecure individuals with CeD and NCGS (n=8)

| Main theme | Corresponding subthemes |
| --- | --- |
| Resources | Costs<br>Time<br>Online resources |
| Social circumstances | Restaurants<br>With friends<br>Travel<br>At home |
| Gluten-free products | Availability<br>Reliability<br>Quality<br>Labeling |
| Mental wellbeing | Personal emotions<br>Social emotions<br>Mental load<br>Coping |

##### Resources

In all cases, participants reported higher costs associated with adhering to a gluten-free diet. Many struggled to quantify the exact amount spent on gluten-free products. One participant commented: *“In a week I eat two to three loaves of bread and a pack or two packs of crackers, so you can easily spend almost €25 to €30 just for the specific gluten-free bread and crackers.” (Participant 7)*.

For seven participants, costs were the reason to buy fewer specific gluten-free products than they would prefer. This included sweet snacks, such as gluten-free cookies, as well as staple products such as gluten-free breads and pasta. For instance, one interviewee stated: *“We have a three-day weekend together and I used to have nice bread every morning. Yes, now I only buy one pack of bread, so I only buy bread for 1.5 days. With snacks, I also try to be more frugal with what I eat. So yes, I now pay more attention to the quantities. Where I would normally eat more from the gluten-free shelf, I now think ’No’.” (Participant 5)*

Other strategies adopted in response to costs included only buying discounted products, baking their own bread, receiving gluten-free food from family, working more hours or implementing energy-saving measures. One participant mentioned indirect cost savings due to the increased difficulty of dining in restaurants and cafes. None of the participants felt driven by financial considerations to consume gluten-containing meals, particularly due to the symptoms they encountered after ingesting gluten.

Regarding the current Dutch diet costs tax compensation, all participants were aware of its existence. While a few participants expressed appreciation for its existence, criticisms of the current system were voiced. This included ambiguity in the process, the annual requirement to provide evidence for CeD, despite it being a lifelong condition, ineligibility for compensation due to various reasons and dissatisfaction with the compensated amount not aligning with the actual expenditure on gluten-free products. In discussing this matter, one interviewee said: *“Well, I don’t get it. I always find it difficult to search every year: ’Hey, where should I be again? What am I supposed to do again?’ So I just think it’s poorly indicated and I also think that a lot of people who have celiac disease don’t know about it.” (Participant 2)*

Only a minority of respondents indicated that the time required to follow a gluten-free diet was a concern, primarily due to time spent reading labels in supermarkets, time spent baking gluten-free products at home or time spent preparing separate meals from family members.

Four participants highlighted the usefulness of online resources, such as gluten-free themed Instagram accounts, gluten-free restaurant apps and general online restaurant information. These resources were found to be helpful for inspiration, staying informed about gluten-free foods and simplifying the process of eating out. One participant commented: *‘Nowadays, of course, many menus are available online. I really like that, because then I can think about ’Okay, that’s a possibility and it comes with that sauce, so 9 times out of 10 that’s not good, maybe they can change that?’” (Participant 8)*

##### Social circumstances

Most participants reported challenges in adhering to a gluten-free diet in social situations. Six participants highlighted difficulties when dining out in restaurants. All of them emphasized the need for pre-research or contacting the restaurants in advance. Five participants noted a lack of knowledge in restaurants, with one expressing: *“But if you were to just go to a restaurant, there is often not much knowledge and nothing at all about cross-contamination.” (Participant 3)*. Limited options in restaurants and menu choices in restaurants also emerged as a recurring issue.

Another concern raised was the constant need to check for cross-contamination handling. Regarding this, one participant shared: *“But I still notice when I walk into a coffee shop or something, sometimes you are lucky that there is an option. But if that option is available, you should also check ’Are there any gluten products standing next to it?’ Or is it a business that understands that gluten-free bread should be kept separate?” (Participant 4)*. There were a few negative experiences shared, where mistakes had been made by the restaurants, even after double checking and pointing out the diet.

In social situations with friends, participants often had to double-check and address mistakes. However, many noted an understanding and benevolence among friends. One participant expressed: *“My friends, they don’t know any better. So when I come for coffee, they always have something sweet for me” (Participant 1)*

Five participants mentioned instances when their gluten-free diet was not being taken seriously unless they explicitly mentioned having celiac disease, rather than just following a gluten-free diet for personal reasons. For example, one said*: “Yes, as long as you don’t mention the disease, they really have the idea that it is just to lose weight and that you would just eat a croquette at home or something.” (Participant 5).* Four individuals linked this perception to the gluten-free diet being a trend of sorts. While two participants acknowledged the trend, they found it to have a positive effect on the overall understanding of gluten-free diets.

All participants mentioned feeling restricted when traveling due to the potential scarcity of gluten-free options in supermarkets and restaurants abroad. The majority addressed this concern by bringing their own gluten-free foods on vacation. As one participant noted: *“Well, we always went camping, but I always had an extra crate of gluten-free food with me, just in case.” (Participant 8).* However, five participants expressed that it was easier to follow a gluten-free diet in some countries than in the Netherlands, citing better options and knowledge, with Italy being mentioned most frequently.

At home, the majority of participants highlighted that their home kitchen situation was well arranged and issues at home were very rare.

##### Gluten-free products

All participants concurred on feeling restricted by the limited availability of gluten-free products in supermarkets. In general, most expressed a desire for more variety in the options. As one participant articulated*: “I think maybe sometimes you miss the choice. When you look at what you could get if you are not gluten-free, you think, oh yeah…” (Participant 3)*.

The majority noted variations in offerings across supermarkets, with some having almost no gluten-free options. Consequently, four participants mentioned deliberately traveling to a more distant supermarket to access a broader selection. Additionally, five participants occasionally ordered gluten-free products online, especially those unavailable in local supermarkets, although they acknowledged this was infrequent due to higher prices. Five participants did mention an increase in gluten-free products compared to previous years.

Four participants raised concerns about many products having ‘traces of’ gluten, making it challenging to find truly gluten-free options. One participant with lactose intolerance also found it difficult to locate gluten-free products without other allergens such as soy or lactose.

Several participants expressed frustration with the unreliability of the gluten-free product offerings. This unreliability appeared in the form of either an inability to rely on the consistent availability of gluten-free products in specific supermarkets or uncertainties about changes in product composition. One individual stated: *It also happens quite often that products suddenly change in composition and that the manufacturer suddenly adds gluten. So that’s annoying too. That is quite often, that the products I always bought suddenly contain gluten. You think, why does that have to happen? (Participant 2)*

Quality concerns also emerged, with three participants expressing dissatisfaction with the taste of some gluten-free products. However, two of them did concur that, overall, they were able to maintain a flavorful diet, while two others recognized a general improvement in taste compared to previous years. Six participants highlighted issues with the healthiness of gluten-free products, citing high sugar content and low fiber as common concerns. One participant explained: *“Because 90% of what is in the store is simply unhealthy. That’s basically the fast food for people with celiac disease. If you go to the store tomorrow, I instruct you to find whole wheat gluten-free bread. You’ll notice it’s just not there. So everyone with celiac disease is almost forced to eat white bread.” (Participant 6)*

Five participants noted that gluten-free or crossed grain logos made shopping easier, eliminating the need to read labels. However, this logo is not consistently accessible in many supermarkets.

##### Mental well-being

There were several prevalent subthemes within the realm of mental well-being that emerged from the interviews. In the personal domain, a recurrent sentiment revolved around feeling on edge or paranoid in eating situations, making it challenging to relax. Expressing this sentiment, one participant remarked: *“You have to constantly pay attention to everything. That’s annoying.” (Participant 5).* Another participant highlighted the struggle by mentioning: *“Sometimes it’s just easier that I think ’Phew, just leave me at home and then I can decide for myself what I eat and…’ Sometimes you just have to pay so much attention and you just don’t always feel like it.” (Participant 8)*

Five participants felt that having to follow a gluten-free diet was a limitation and/or difficulty. One interviewee, when asked about how they experienced having to follow a gluten-free diet, answered: *“Like a prison” (Participant 6).* However, contrasting views were present, with another participant expressing a positive outlook due to the experienced relief of symptoms.

In terms of feelings towards others, six participants revealed a common sentiment of feeling like a burden to those around them. As one participant put it: *“Because I always feel a little uncomfortable towards other people. When I eat with others or in restaurants, they always have to do a lot more for me. In addition, you also have to check a bit whether it truly went well, so I always feel very embarrassed towards other people and I especially find that very sad.”* (Participant 5)

Another recurring theme was the mental load associated with adhering to the gluten-free diet, requiring constant thought about food-related considerations. One participant illustrated this, saying: *“For example, if I go somewhere, to another city or whatever, I have to think carefully about: ’How long will I be there? Should I take something with me? Or should I just eat beforehand and be fine? Or are there any shops nearby that I know I can go to? ’I always have to think about that.”* (Participant 4)

A prevalent sentiment throughout the interviews was the acceptance of the limitations that accompany following a gluten-free diet. Participants described this acceptance as ’being used to it,’ ’getting over it,’ or ’putting it in perspective.’ One participant stated: *“Well, I’ve been doing it for a long time now, so you’re already used to a lot of things and you know how it works, so to speak.”* (Participant 3). Another participant noted: *“For example, my sister and my brother, both of them don’t have it [celiac disease], so I’m always jealous of that. Well, I’d like to trade for a day, wouldn’t I? But hey, I always say, there are always worse things. And at some point, you have such an understanding and know what it is all about…”* (Participant 1)

## DISCUSSION

This study aimed to characterize food insecurity among Dutch individuals with CeD and NCGS, revealing that a substantial proportion (23%) was food insecure, with 10% facing very low food security. Food insecurity was associated with younger age, having a lower income and lower educational level, higher perceived financial scarcity and poorer mental well-being. Furthermore, very low food insecurity was associated with a lower diet quality score. Our comprehensive exploration of perceived barriers to adhering to a gluten-free diet, through both quantitative and qualitative approaches, exposes multifaceted challenges in food insecure individuals across several domains, including resources, social circumstances, gluten-free products and mental well-being.

The prevalence of food insecurity in our study was similar to previous reported data from the United States among individuals with CeD (23, 24), and higher than previous reported data among a general sample of adults with a relatively low socioeconomic position living in the Netherlands (33). Our findings confirm the established correlation between food insecurity, lower income and educational level(34). Another observation is that participants experiencing food insecurity tended to be younger, possibly due to limited financial resources and potentially less stable employment. Additionally, an association between food insecurity and perceived financial scarcity was identified, which highlights financial difficulties that food insecure participants may have in other aspects of their lives. Recognizing the potential combined impact of financial scarcity and food insecurity is crucial, as previous research has linked financial difficulties to adverse health outcomes and diminished overall well-being(35).

Results of our study revealed an association between a lower diet quality and food insecurity. This finding aligns with previous studies that also identified a connection between food insecurity and diet quality(36–38), extending to individuals following a gluten-free diet (24).The observation that only those experiencing very low food security exhibited a lower diet quality in this study, suggests that there may be a nuanced relationship between different levels of food security and diet quality, with very low food security having a more pronounced impact. Notably, unlike previous studies on food insecurity(39–41), we found no association between food insecurity and BMI. This indicates potential variations in the impact of food insecurity on body weight across different populations.

Another possible explanation for the absence of this association could be the gluten-free diet limiting food choices, potentially mitigating the effects of food insecurity on BMI. Additionally, individuals with CeD typically have lower BMIs compared to the general population(42), which could also contribute to the lack of association observed in our study.

The study sheds light on the mental and emotional challenges associated with adhering to a gluten-free diet, revealing feelings of restriction, burden, and the need for constant vigilance. Our research indicated a lower self-reported mental wellbeing in all participants with CeD and NCGS compared to the average Dutch population, based on the most recent data available from 2016(43), aligning with previous studies emphasizing the impact of adhering to a gluten-free diet on mental health(44, 45). Particularly noteworthy, self-reported mental wellbeing was even lower among food insecure individuals, echoing results from a prior study reporting lower reported mental health among food insecure individuals on a gluten-free diet(46). This underscores the potential contribution of food insecurity to an increased risk of worsened mental health in people with CeD and NCGS.

Building upon previous research that identified common barriers into adhering to a gluten-free diet(47–49), our results emphasize heightened barriers for food insecure individuals across various areas. Unsurprisingly, the availability of resources was a significant experienced barrier in adhering to the gluten-free diet. Moreover, our exploration revealed intensified challenges extending beyond economic constraints, including social circumstances and aspects related to gluten-free products.

Although social circumstances and naturally gluten-free products did not retain statistical significance in the adjusted model for individuals with very low food security, the persistent trend suggests that the lack of significance might be attributed to statistical considerations, concerning subgroup size. While our survey did not assess gluten-free diet adherence, previous research reported an increase in intentional gluten ingestion among individuals experiencing food insecurity(24). Additionally, research suggests that adherence might be related to perceived barriers(50, 51). Though none of the participants in our interviews mentioned deliberately eating gluten, it is important to recognize that our study had a limited number of participants, and thus, the absence of such reports may not fully represent the broader population. Additionally the established correlation between gluten ingestion and reduced nutritional uptake (25) could be exacerbated by our observation that individuals experiencing food insecurity tend to have a lower diet quality. Given its implications for nutritional status and overall health outcomes, intentional gluten ingestion among food insecure people with CeD remains a possible area of concern.

Several strengths of this study can be acknowledged. Firstly, it offers previously unexplored insights into food insecurity among individuals with CeD and NCGS in the Netherlands. Secondly, by employing a mixed-methods study design, we gained further insight into understanding the challenges individuals face. However, it is important to also recognize limitations of the current study. First, the cross-sectional design of our study makes it impossible to draw causal conclusions regarding factors associated with food insecurity. However, it is more plausible that food insecurity lies within the causal pathway of diet quality and perceived barriers rather than the reverse. Second, this study was based on self-reported data through anonymous surveys, which introduces the possibility of recall bias and social desirability bias, leading to potential inaccuracies in the data.

Additionally, the reliance on self-reported diagnoses of celiac disease and non-celiac gluten sensitivity may lack the accuracy of clinical confirmation. Third, the study participants were recruited through the Dutch Celiac Association, thus the characteristics of the study population might not be reflective of the larger population of people with CeD and NCGS in the Netherlands. Previous findings linked membership of celiac disease advocacy groups with improved gluten-free diet adherence, suggesting a potential bias towards a more health-conscious sample(52). However, given the nature of the online survey research, alternate options for recruitment were essentially limited. Fourth, the gender distribution exhibited a significant imbalance (90% female), which can be attributed in part to the higher prevalence of CeD in females (53) and the pre-existing disparity in the Dutch Celiac Association database, where 70% are female (personal communication Dutch Celiac Association, January 2024). Additionally, this skew may be influenced by potential differences in willingness to participate in such surveys between genders(54). Finally, it should be noted that parts of the questionnaire were not previously validated, such as the addition of ‘gluten-free’ to the USDA 6-item food security scale. Thus, the interpretation of responses regarding food insecurity might be limited to the concept of ‘gluten-free food insecurity’, which has been previously described in otherwise food-secure households(23). This perception was reinforced during the interviews, as most participants acknowledged having sufficient funds for food in general, yet encountered challenges in affording specific gluten-free options.

## Conclusions and implications

In conclusion, our findings reveal that food insecurity is prevalent among people with CeD and NCGS in the Netherlands and indicate an association with poorer diet quality and higher perceived barriers to adhering to a gluten-free diet. The exploration of perceived barriers provides deeper understanding of the specific challenges faced by food insecure individuals with CeD and NCGS. Clinicians should take these challenges into account in their recommendations to individuals with CeD and NCGS that may experience food insecurity. Furthermore, insights from this study underscore the need for public initiatives to address the multifaceted impact of food insecurity in this population and may inform policy makers to develop targeted interventions to improve access to gluten-free foods. Future research should focus on enhancing our understanding of these complex dynamics to inform evidence-based interventions for improving access to gluten-free diets for food insecure individuals with CeD and NCGS.

## STATEMENTS

### Contributors

SS: conceptualized the study, conducted the interviews, performed the data analyses, drafted the initial manuscript, reviewed and revised the manuscript. LV: conceived and conceptualized the study, supervised data analyses, reviewed and revised the manuscript. JK: scientific project leader and guarantor, conceived the study, reviewed and revised the manuscript. All authors approved the final manuscript as submitted.

### Data availability statement

Data are available upon reasonable request.

